# PATIENT-ASSESSED QUALITY OF CARE AND LEVEL OF SATISFACTION IN HEALTHCARE FACILITIES IN CROSS RIVER STATE, NIGERIA

**DOI:** 10.1101/2023.07.12.23292492

**Authors:** John John Etim, Glory M. Nja, Regina Idu Ejemot-Nwadiaro

## Abstract

**Background:** Patients’ experiences with health care professionals are a central component of quality of care, complementing more technical aspects of care such as the appropriate use of medications and procedures.

**Aims:** This study sought to assess patients’ quality of care and level of satisfaction in healthcare facilities in Cross River State, Nigeria.

**Methods:** The study adopted a descriptive cross sectional survey employing quantitative data collection method that utilized a semi-structured questionnaire to collect data from patients. A multi-stage sampling procedure was used where 405 respondents were selected through simple random sampling. A 27 items semi-structured questionnaire was used to elicit data from respondents. Data collected were analyzed and presented using descriptive statistics.

**Results:** On a multi response options, the study showed that more than half 294(72.8%) respondents were satisfied with the services provided by healthcare professionals on their current admission; more than half of the respondents 235(58.2%) were satisfied with previous care received; about half 197(48.8%) of the respondents complained about long waiting time to be seen on admission; more than half 209(51.7%) of the respondents strongly disagreed that the hospital/ward environment into which they were admitted was clean and conducive, with a significant proportion 335(82.9%) who were unable to get all the prescribed drugs from the facility, while just 69(17.2%) of those not satisfied could recall some instances/aspects of healthcare they were not satisfied with where 49(12.2%) recalled poor attitude of healthcare professionals.

**Conclusions:** Timely care and better communication between patient and healthcare providers were recommended among others.

**What is already known on this topic:**

- There is a recorded level of patients’ dissatisfaction with healthcare in the study setting.

**What this study adds:**

- Poor attitude of healthcare workers has been associated with patients’ dissatisfaction with healthcare.
- Poor attitude of healthcare workers has been associated with poor work environment which by extension affect the mental health of healthcare staff.

**How this study might affect research, practice or policy:**

- This study outlines the implication of poor work environment on mental health of workers as by extension affects patients’ quality of care and as such calls better healthcare management by trained experts.

## Introduction

Patients-assessed quality of care is a reflection of patients’ perspective on meeting the needs of a client who visits health facilities in line with their expectations and patients’ contentment with the way and manner health services are provided to them. Patients’ experiences with health care professionals are a central component of quality of care, complementing more technical aspects of care such as the appropriate use of medications and procedures. Patient-assessed quality of care looks at the dimensions of quality in healthcare necessary for adequate and appropriate patient care which includes: technical competence, access to services, efficiency, interpersonal relations, continuity, safety, amenities. Hence quality of care and healthcare outcomes can be measured by assessing whether health care is effective, accountable, safe, fair and accessible to patients in order to influence their perspective and opinion concerning the quality of care being received. To fulfil these dimensions, there are some factors that are essential determinants of patients’ perception of quality of care and satisfaction when health facilities are visited, these include: the attitude of healthcare providers, sanitation (cleanliness of the hospital environment), availability of prescribed drugs, availability of basic infrastructures like constant power supply (electricity), water supply, couch in outpatient department, etc. and the waiting time (promptness of service that translates the agility of healthcare professionals).^1^

A study in 2014 used patients to assess the quality of care among healthcare providers and reported that among all the respondents assessed, 80.1% were found to be satisfied (those who reported to be very satisfied and satisfied) with the outpatient services of the hospital whereas the remaining 19.9% were dissatisfied.^2^ In the same study, level of satisfaction by fully exhausted scales showed that 11.7, 68.4, 2.4, 12.2, and 5.3% of respondents reported to be very satisfied, satisfied, neutral, dissatisfied and very dissatisfied respectively.^2^ Binary and multiple logistic regressions were performed to identify factors associated with patients’ satisfaction using different covariates where respondents who claimed to have had a long stay in the hospital were found to be more satisfied than those who claimed to have had a very long stay.^2^ On the other hand, respondents who did not get all the required items/services from the hospital were less satisfied than their counterparts. Absence of good dialogue with outpatient service providers was also found to be negatively associated with respondents’ satisfaction. The results of their study showed that 80.1% of patients were satisfied with the outpatient health service they received.^2^ Patients’ satisfaction was associated with length of stay to receive care (waiting time), presence of good dialogue with service providers, maintenance of privacy during care, the favourability of situations to ask questions, and availability of required services.^2^

Attitude of healthcare providers is made manifest in the level of communication between patients and healthcare providers. Communication is a critical skill in healthcare delivery. It is a dynamic process used to gather assessment data, teach, persuade, and express caring and comfort from patients. It is an integral part of the healthcare professionals–patient relationship. A study carried out in Chicago on the correlation between patients’ comprehension of their reason for hospital admission and overall patients’ satisfaction revealed that effective communication between doctors and patients about the reason for their admission can be an important predictor of patients’ satisfaction and measure of quality of care.^3^

A study in India reported that overall level of satisfaction with doctors ranged from 89.29% to 9.96%, where satisfaction with the attitude of nurses was slightly lower than 9.96%.^4^ A study in Pakistan showed that 48.8% of the patients were not satisfied with the patient doctor communication and 27.6% said they did not receive helpful support from the nurses.^5^ A study done in Iran revealed that 46.2% of patients were satisfied with admission services of the hospital, the highest score in the satisfaction areas was in the areas of attitude of staff.^6^ A study conducted in a tertiary hospital in Dhaka revealed that physical evidence, doctor’s services, nurse’s services and feedback from patient lead to a higher level of patient satisfaction, and that amongst these variables, doctors service orientation was the most important factor explaining patient satisfaction.^7^ In an Ethiopian study, a scale (very politely, politely, neither politely nor impolitely, impolitely, and very impolitely) was used to assess the degree of politeness/impoliteness of outpatient service providers who served patients, 15.7% and 69.7% of respondents described outpatient service providers as very polite and polite during service provision while the remaining 10.8%, 3.1% and 0.7% of respondents described outpatient service providers as neither polite nor impolite, impolite, and very impolite, respectively.^2^

Prolonged waiting time can cause the patient to lose his/her confidence on the whole healthcare process in any health facility. An earlier study in Nnamdi Azikiwe University Teaching Hospital (NAUTH) reported that 79% of patients were satisfied with services in the general out-patient department in terms of the availability, accessibility, convenience and manner of delivery by hospital personnel.^8^ It also identified areas in the service delivery that needed to improve such as the comfort of the waiting area, process of retrieving records and availability of some medicine within the hospital pharmacy and at a cheaper rate.^8^ Respondents were asked to estimate the amount of time they spent to use various hospital services to determine the total waiting time in a study conducted among 35.2% and 31.9% who visited medical and surgical outpatient departments of the Hawassa University Teaching Hospital in Southern Ethiopia, respectively, nearly one-third (32.7%) of patients waited for more than 90 minutes to enter to the outpatient departments after they have gone through registration process, among the total respondents, 50.5% had a length of stay of more than two hours in the hospital and based on the respondents’ rating of the length of stay they had to receive care, the length of stay was reported to be very long by more than one-fourth (26.3%) of respondents.^2^

The absence of prescribed drugs can be very frustrating to the patient and even the healthcare professionals. Patients also use this dimension to measure the quality of care received. It was documented in a study that 71.2% reported to have got all ordered services from the hospital (laboratory tests, diagnostic services and prescribed drugs).^2^ An earlier study in Nnamdi Azikiwe University Teaching Hospital (NAUTH), reported that 79% of patients were satisfied with services in the general out-patient department in terms of the availability, accessibility, convenience and manner of delivery by hospital personnel.^8^ It also identified areas in the service delivery that needed to improve such as the comfort of the waiting area, process of retrieving records and availability of some medicine within the hospital pharmacy and at a cheaper rate.^8^ In Pakistan, respondents registered their bitterness in respect to the absence of prescribed drugs and they submitted that the absence of the drug in the health facility has caused them more pain than gain as they found themselves paying more than they ought to pay if they were to get these drugs in the facility.^5^ They as well reported that the efficacy of drugs bought from outside the facility is always in doubt.^5^ Furthermore, going out to get drugs will sometimes delay the process of treatment and sometimes contribute to complications or patient’s death.^5^ It is in this light that a relationship between availability of prescribed drugs and patient’s satisfaction with quality of care was wsterblished.^4^

The aesthetics of a hospital environment gives the patient hope of coming out alive from the health facility. This also contributes to patient’s satisfaction of quality of care. A study carried out on patients’ perception of obstetric practice in Calabar, Nigeria reported that poor sanitary condition and lack of basic amenities were the major cause of patients’ dissatisfaction.^9^ The area of cleanliness of the hospital environment had the lowest score for satisfaction as only 33% were satisfied with sanitation in an Indian study which affected the overall satisfaction of outpatients.^4^ It was concluded in that study that sanitation is an essential factor in patients’ satisfaction.^4^ A study conducted to investigate patients’ satisfaction with services obtained from Aminu Kano Teaching Hospital, Kano, Northern Nigeria showed that patients complained that the environment of the hospital gives them hope of returning home alive after been admitted for treatment in the facility.^10^ The study also found that patients registered their displeasure of how the outpatient clinics were not hygienic and that, such unhygienic hospital environment can cause nosocomial infections to patients order than recovering from the ailment that brought them to the hospital.^10^ A study on Patient satisfaction on admission in Nnamdi Azikiwe University Teaching Hospital, Nnewi, Nigeria also found a similar scenario as patients tied their satisfaction to the sanitation of the environment.^1^

The level of patients’ satisfaction is one among the mechanisms used in assessing the quality of health care services and addressing patients’ expectations was found to be associated with high client satisfaction and better health outcomes.^11^ A study carried out on patients’ satisfaction with services at Aminu Kano Teaching Hospital, Kano, Nigeria revealed that 88%, 87% and 84% of the patients were satisfied with patient-provider relationship, inpatient services, hospital facilities and access to care respectively.^10^ A study to find out the correlation between patient comprehension of their reason for hospital admission and overall patient satisfaction in the emergency department and found out that power failure, poor infrastructure, absence of water supply and other basic hospital amenities can determine patience satisfaction regarding the healthcare services received.^3^ Their study found a correlation between these variables researched on.^3^ Similarly, a study found a relationship between the availability of quality hospital equipment and the satisfaction of patients on the quality of care received in Teaching Hospital in Anambra State, Nigeria.^8^ However, this present study assessed patients’ quality of care and level of satisfaction in healthcare facilities in Cross River State, Nigeria.

## Methods

### Study area

This study was conducted in Central Senatorial District of Cross River State which stretches between longitude 7.00 to 858E and Latitude 5.00 to 5.29N. The area is located midway between Cross River Southern and the Northern Senatorial Districts and bounded in the north by Ogoja Local Government Area and in the south by Biase Local Government Area. The Republic of Cameroon and Ebonyi State form the Eastern and Western boundaries respectively. It has a projected 2017 population of 1,163,903 (590,527 Males and 573,376 Females) from the 2006 census figures and it occupies an area of 2005 square Kilometres. According to the statistics of Cross River State Ministry of Health (2018), there are six (6) government secondary health facilities, forty-one (41) private health facilities and one hundred and thirty three (133) Primary Healthcare Centers that provide healthcare services to patients in Central Senatorial District of Cross River State. That is, there are a total of one hundred and eighty (180) healthcare facilities in the study area.

### Study design

The study adopted a descriptive cross sectional survey employing quantitative data collection method that utilized a semi-structured questionnaire to collect data from patients.

### Study population

The population of the study comprised all patients admitted in the selected healthcare facilities (primary, secondary and private facilities) at the time of the study.

### Sample size determination

The formula below was used to determine the sample size for patients.

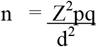

where;

n = Sample size

Z = Z-score which is 1.96 at 95% confidence limit

p = prevalence or probability of the event occurring (71.5% = 0.712)

*P was the prevalence of patients’ satisfaction of healthcare delivery from previous study.^2^

q = probability of the event not occurring (1-p = 1-0.712 = 0.287)

d = acceptable margin of error (5% = 0.05)

Therefore,

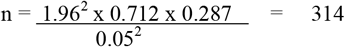

Calculating for 29% non-response rate 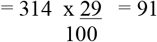

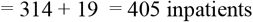

### Sampling procedure

A multi-stage sampling procedure was used as follows:

#### Stage-I: Selection of Senatorial District

Through convenience sampling technique, Central Senatorial District was selected out of the three (3) Senatorial Districts (Northern, Central, and Southern) in Cross River State based on the fact that such study has not been found in the District. Figure-1.

**Figure-1.**
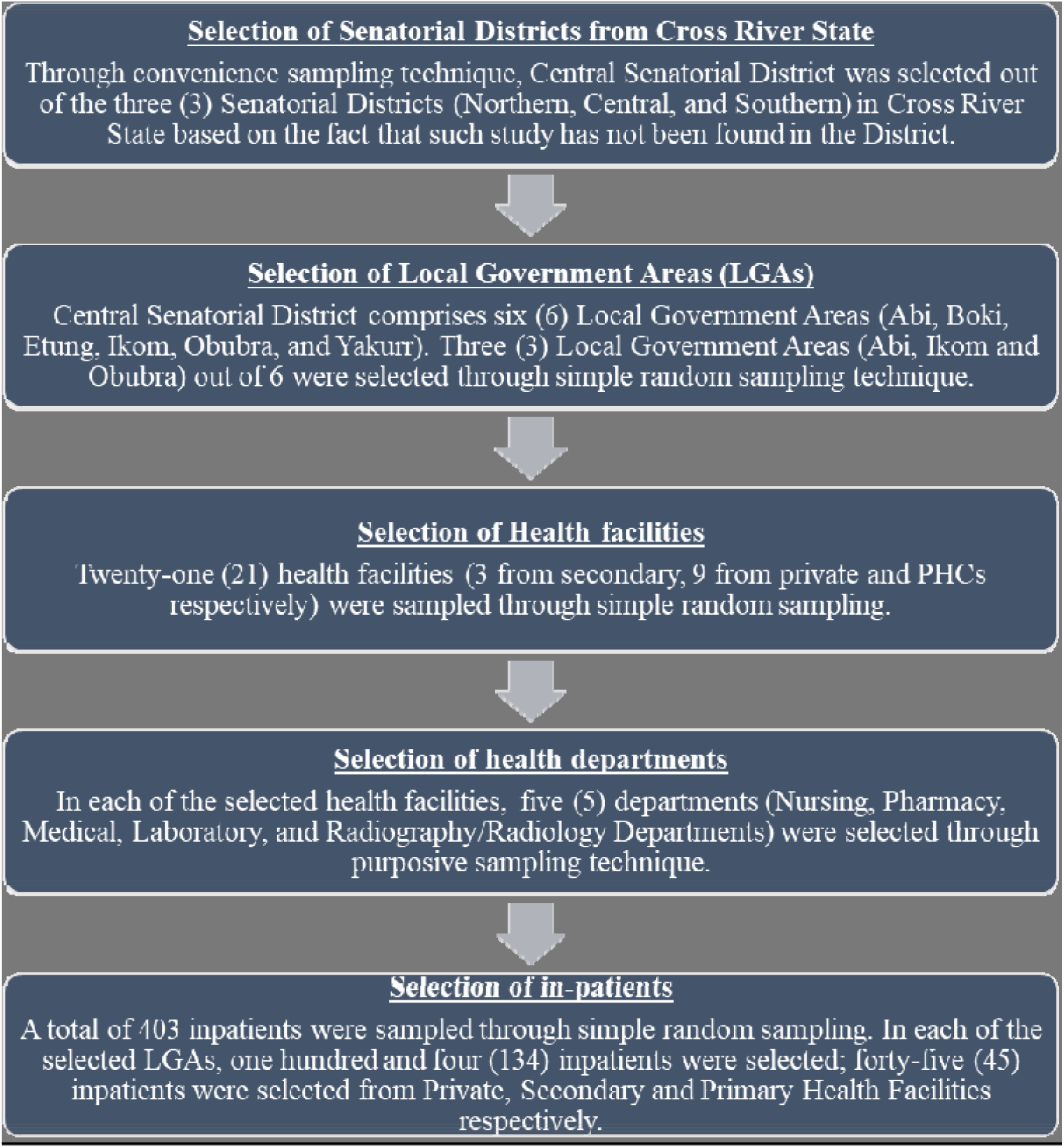
Flow-chat showing the sampling procedure in the study

#### Stage-II: Selection of Local Government Areas

Central Senatorial District comprises six (6) Local Government Areas (Abi, Boki, Etung, Ikom, Obubra, and Yakurr). Three (3) Local Government Areas out of 6 were selected through simple random sampling technique. This was done by writing out the names of the six (6) Local Government Areas in the Central Senatorial District on pieces of papers, folded into a container and thoroughly mixed, from which three LGAs were drawn one after the other without replacement. Through this method, Abi, Ikom and Obubra LGAs were selected for the study respectively. Figure-1.

#### Stage-III: Selection of healthcare facilities

The names of all the health facilities in each of the selected LGAs were written on pieces of paper and dropped in three (3) separate containers clearly marked Private Health Facilities, Secondary Health Facilities and Primary Health Facilities. These were thoroughly mixed, thereafter three (3) pieces of papers were selected from the container marked Private Health Facilities; One (1) from Secondary Health Facilities and three (3) from the Primary Health Facilities. This process was repeated in the remaining two (2) LGAs, yielding a total of twenty-one (21) health facilities; nine (9) from the Private Health Facilities, three (3) from the Secondary Health Facilities and nine (9) from Primary Health Facilities. Figure-1.

#### Stage-IV: Selection of departments

In each of the selected health facilities, five (5) departments (Nursing, Pharmacy, Medical, Laboratory, and Radiography/Radiology Departments) were selected through purposive sampling technique. Figure-1.

#### Stage-V: Selection of in-patients

A total of four hundred and three (403) inpatients were sampled through simple random sampling. The choice of this sampling technique was to allow the researcher collect data from inpatients who were conscious and willing to partake in the study without introducing bias. Numbers were assigned to patients from each ward that guided the sampling. In each of the selected LGAs, one hundred and four (134) inpatients were selected; forty-five (45) inpatients were selected from Private, Secondary and Primary Health Facilities respectively. Figure-1.

### Instruments for data collection

A semi-structured questionnaire was prepared based on the research objectives formulated for inpatients. The questionnaire for inpatients consisted of three (3) sections (A, B, and C) with a total of 27 questions. Section A had 5 questions to elicit information on the socio-demographic characteristics of patients; Section B had 9 questions to elicit information on accessibility to healthcare services; and Section C had 13 questions to elicit information on patient-assessed quality of care.

### Pre-testing of research instrument

Forty (40) copies of the questionnaire representing 10% of the sample size for in-patients were pretested among 40 inpatients in the Southern Senatorial District of Cross River State. This was done to test for reliability and validity of the instrument. This was to find out if there is any consistency in the items of the instrument and to ascertain the suitability of the instrument for the study. Data from the respondents were collected once and were coded, scored, and analyzed. The instrument administered were subjected to reliability test using Cronbach Alpha reliability analysis to determine the reliability estimate of the instrument. The reliability index ranged from 0.80 – 0.97. Cronbach Alpha reliability estimate of the test instrument was done in order to determine the internal consistency of the instrument items. These values were considered high enough to be used for the study.

### Methods of data analysis

Data collected were coded, entered and analyzed using Statistical Package for Social Sciences (SPSS) software (version 20.0). Results computed were expressed in simple percentages and presented in tables and charts.

## Results

### Socio-demographic characteristics of patients

A total of 403 in-patients participated in the study giving a response rate of 99.5% with most 339(84.0%) females and 64(16.0%) males. A greater proportion 121(30%) of the respondents were within the age bracket 23-27years, followed by those 79(19.6%) within the age bracket 28-32years and 66(16.3%) within the age bracket 18-22years. The mean age of the respondents was 29years. Most of the respondents 240(59.4%) were married, with 134(33.2%) single. Almost all respondents 400(99.0%) were Christians with greater proportion 87(21.5%) Catholics, followed by 76(18.8%) Redeemers and 65(16.1%) Apostolic, respectively (Table 1).

**TABLE 1.** Socio-demographic characteristics of inpatients

| **Variables** | **Frequency(n=403)** | **Percentage (%)** |
| --- | --- | --- |
| **Gender** |  |  |
| Male | 64 | 16.0 |
| Female | 339 | 84.0 |
| **Age (in years)** |  |  |
| 18-22 | 66 | 16.0 |
| 23-27 | 121 | 30.0 |
| 28-32 | 79 | 20.0 |
| 33-37 | 40 | 10.0 |
| 38-42 | 49 | 12.0 |
| 43-47 | 29 | 7.0 |
| 48 and above | 19 | 5.0 |
| **Marital status** |  |  |
| Single | 134 | 33.2 |
| Married | 240 | 59.4 |
| Divorced | 8 | 2.0 |
| Widowed | 12 | 3.2 |
| Separated | 9 | 2.2 |

### Accessibility to healthcare services

More than half 303(75.0%) of respondents had ever visited a health facility on account of ill health; of this, majority 133(32.9%) visited general hospitals, followed by 87(21.5%) who visited private hospitals, while 85(21.0%) visited PHCs. A greater proportion 139(34.4%) stayed 1-3days on admission, while 26(6.4%) stayed 13-15days on admission. Most of the respondents 134(33.2%) were admitted into female medical ward with the least 8(2.0%) admitted into male surgical and paediatric ward wards respectively. More than half of the respondents 235(58.2%) were satisfied with previous care received, while just 69(17.2%) of those not satisfied could recall some instances/aspects of healthcare they were not satisfied with where 49(12.2%) recalled poor attitude of healthcare professionals (Table 2).

**TABLE 2.** Patient-assessed quality of health care delivery

| Assertions/statements | Freq.(N) | (%) |
| --- | --- | --- |
| **Ever visited health facility before the present visit** |  |  |
| Visited | 302 | 75.0 |
| Never visited | 101 | 25.0 |
| **Total** | **403** | **100** |
| **Health facility ever visited by patient before the present visit** |  |  |
| Private hospital | 87 | 21.5 |
| General hospital | 133 | 32.9 |
| PHC | 85 | 21.0 |
| **Total** | **305** | **75.5** |
| **Length of stay during the visit to the health facility** |  |  |
| 1-3days | 139 | 34.4 |
| 4-6days | 65 | 16.1 |
| 7-9days | 40 | 9.9 |
| 10-12days | 35 | 8.7 |
| 13-15days | 26 | 6.4 |
| **Total** | **305** | **75.5** |
| **Ward admitted into during the last visit** |  |  |
| Male Medical | 26 | 6.4 |
| Male Surgical | 8 | 2.0 |
| Male Orthopaedic | 9 | 2.2 |
| Female Medical | 134 | 33.2 |
| Female Surgical | 19 | 4.7 |
| Female Orthopaedic | 12 | 3.0 |
| Maternity | 45 | 11.1 |
| Gynaecology | 13 | 3.2 |
| Accident/Emergency | 31 | 7.7 |
| Paediatric | 8 | 2.0 |
| **Total** | **305** | **75.5** |
| **Satisfaction level on the past visit to health facility** |  |  |
| Satisfied | 235 | 58.2 |
| Not satisfied | 70 | 17.3 |
| **Total** | **305** | **75.5** |
| **Those who can remember why not been satisfied on last visit** |  |  |
| Can remember | 69 | 17.2 |
| Cannot remember | 1 | 0.2 |
| **Total** | **70** | **17.4** |
| **Reasons why patients were not satisfied with services on their previous visit** |  |  |
| High cost/bills | 20 | 4.99 |
| Poor attitude of healthcare professionals | 49 | 12.2 |
| **Total** | **69** | **17.2** |

Majority of the respondents 138(34.2%) were on current admission into female medical ward, followed by 74(18.3%) admitted into maternity ward, with least admission into male surgical 11(2.7%). A greater number 160(39.6%) spent 1-3days on current admission with a lesser proportion 11(2.7%) who have spent one month (Table 3).

**TABLE 3.**
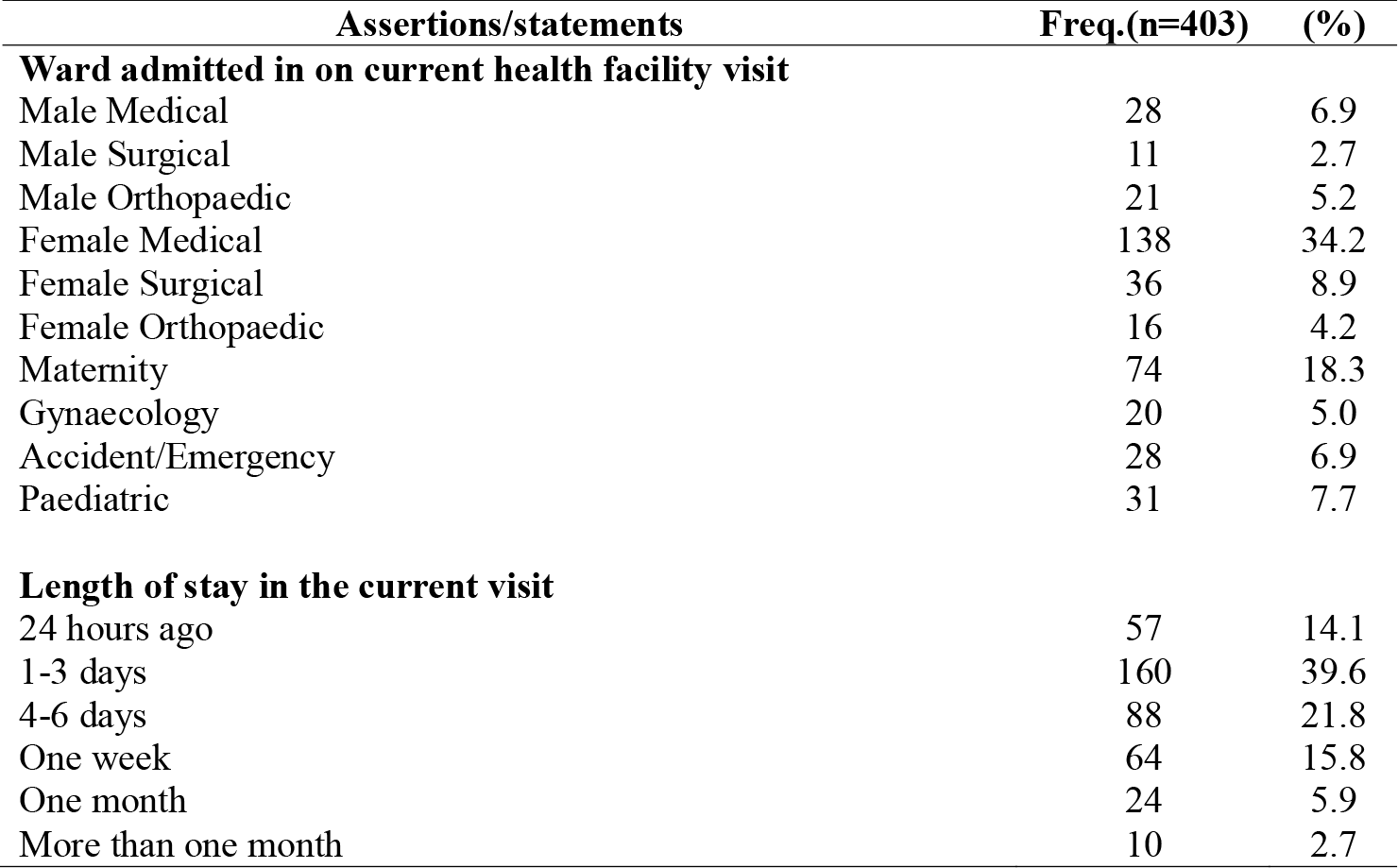
Patients’ accessibility to healthcare services and length of stay on current admission

| **Assertions/statements** | **Freq.(n=403)** | **(%)** |
| --- | --- | --- |
| **Ward admitted in on current health facility visit** |  |  |
| Male Medical | 28 | 6.9 |
| Male Surgical | 11 | 2.7 |
| Male Orthopaedic | 21 | 5.2 |
| Female Medical | 138 | 34.2 |
| Female Surgical | 36 | 8.9 |
| Female Orthopaedic | 16 | 4.2 |
| Maternity | 74 | 18.3 |
| Gynaecology | 20 | 5.0 |
| Accident/Emergency | 28 | 6.9 |
| Paediatric | 31 | 7.7 |
| **Length of stay in the current visit** |  |  |
| 24 hours ago | 57 | 14.1 |
| 1-3 days | 160 | 39.6 |
| 4-6 days | 88 | 21.8 |
| One week | 64 | 15.8 |
| One month | 24 | 5.9 |
| More than one month | 10 | 2.7 |

### Patient-assessed quality of care

A total of twelve items in the questionnaire were used to measure patient-assessed quality of care where more than half 294(72.8%) respondents were satisfied with the services provided by healthcare professionals on their current admission; about half 197(48.8%) of the respondents complained about long waiting time to be seen on admission; more than half 209(51.7%) of the respondents strongly disagreed that the hospital/ward environment into which they were admitted was clean and conducive, with a significant proportion 335(82.9%) who were unable to get all the prescribed drugs from the facility. Scores were assigned to responses related to this section of the questionnaire which were later summed up to get the total for each individual and mean score calculated to be 36. All scores range less than 36 represented responses for poor quality of care, all scores range that sum up to 36 represented responses for moderate quality of care, while all scores range greater than 36 represented responses for high quality of care respectively. The minimum recorded score was 1 while the maximum recorded score was 48 out of a possible total of 48. The score of all the respondents was then collated (Table 4).

**TABLE 4.** Patient-assessed quality of care

| Quality of care | Range (Minimum score =1;<br>Maximum score = 48) | Frequency<br>(N) | Percent<br>(%) | Mean score |
| --- | --- | --- | --- | --- |
| **Poor** | Summed scores less than mean score | 130 | 32.3 | **36.0** |
| **Moderate** | Summed scores equal to mean score | 15 | 3.7 |  |
| **High** | Summed scores greater than mean score | 258 | 64.0 |  |
| **Total** | **48** | **403** | **100** |  |

## Discussion

### Main findings on patient-assessed quality of care and level of satisfaction

A total of 403 inpatients (16.0% males and 84.0% females) were used for the study. Analysis on the data collected from inpatients showed that 64% were satisfied with the services provided by healthcare professionals. This finding is similar with a study that used patients to assess the quality of care among healthcare providers and reported that among all the respondents assessed, 80.1% were found to be satisfied with the services.^2^ To further support this, a study in India 89.29%, Pakistan 51.2%, and Iran 46.2% of patients were satisfied with services of the health staff.^4,5,6^ Furthermore, a study in Nigeria reported 78% of patients’ satisfaction with healthcare services in Nnamdi Azikiwe University Teaching Hospital (NAUTH).^8^

The study finding further showed that half of the patients (50%) complained of prolonged waiting. This is in line with the finding of a study where 48.8% patients complained of waiting for too long in hospital.^2^ About 3.5% complained of not been involved and informed in decisions about their care and healthcare professionals not been polite (3.2%), and not explaining the treatment/health advice in a way they could understand (4.2%). To support this finding, a study reported that 3.1% complained of healthcare professionals being very impolite.^2^ A total of 52.2% registered their displeasure with hospital/ward environment into which they are admitted being unclean and not conducive. To support this finding, it was reported that 67% of patients complained of unclean hospital environment.^9^ Similarly, a study found out that patients registered their displeasure of how the clinics were not hygienic and that, such unhygienic hospital environment can cause nosocomial infections to patients order than recovering from the ailment that brought them to the hospital.^10^ In addition, almost all the patients (82.9%) complained of not being able to get all the prescribed drugs in the facility. To corroborate this, 71.2% and 79% was reported in previous studies as patients complained same.^2,8^ Despite all these complains from patients, 88.8% were confident in the healthcare professionals’ ability to treat their illness, and are ready to recommend the healthcare facility and the services to their family and friends. This is not different from the finding in a previous study where 88% patients were confident in caregivers, 87% believing the treatment and 84% being ready to recommend the services.^10^

Conclusively, some 55(43.8%) of the patients who rated the serviced received being poor were not satisfied with the quality. Long waiting time, unclean healthcare hospital/ward environment, inability to get all the prescribed drugs from the facility, and poor attitude of healthcare professional resulting from poor work environment as source of stress and depression to workers were some factors identified to influence patients’ dissatisfaction with healthcare delivery. The following recommendations are made based on the key findings from this study: Patients care should be provided in clean environment for better health care as this will improve patients’ confidence on healthcare professionals and the facility respectively; There should be competent healthcare providers in healthcare facilities; There should be availability of consumables and frequent electricity; Swift response to patient to reduce prolonged waiting time and improve care that is timely is recommended, and; Healthcare professionals are to communicate professionally and respectfully to patients.

### Limitations

This study could be limited by study design and sample size which may affect generalizability of the study finding. The researchers were able to solve this limitation by making inference based on the findings of the study and limiting its conclusion based on the sample used in the study.

## Data Availability

Data can be produce on request

## Acknowledgements

The dedicated effort of Dr. Glory Nja in ensuring that this work attains some level of quality is well appreciated. The authors appreciate the effort of all post-graduate staff of Department of Public Health, Faculty of Allied Medical Sciences, College of Medical Sciences, University of Calabar, Nigeria. Finally, the participants of this study are well appreciated.

## Ethical approval

Ethical clearance was obtained from Cross River State Research Ethics Committee with the Rec. No. CRSMOH/RP/REC/2018/119. Permission for data collection was obtained from heads of each facility/departments used in the study. During data collection, verbal informed consent was sought from respondents, upholding the confidentiality of their responses and protection, including their voluntary participation in the study. Respondents were aware of the freedom to withdraw from the study at any time if they perceive discomfort resulting from the study. The significance and benefits of the study was explained to the understanding of the respondents. To ensure their confidentiality and maintain anonymity, respondent’s responses were not disclosed to any third party especially the facility management and no name was written on the instrument for data collection. Data generated is stored electronically and will be destroyed after five years.

